# Prognostic value of compact myocardial thinning in patients with left ventricular non-compaction

**DOI:** 10.1101/2024.12.19.24319385

**Authors:** Tea Gegenava, Martijn Tukker, Kadir Caliskan, Alexander Hirsch, Ashish Manohar, Seung-Pyo Lee, Anjali Owens, Deborah H. Kwon, Jay Ramchand, Matthew T. Wheeler, W. H. Wilson Tang, Koen Nieman

## Abstract

**Introduction:** Left ventricular non-compaction (LVNC) can be associated with a wide spectrum of clinical presentations, ranging from asymptomatic to ventricular tachyarrhythmia (VT), heart failure (HF), and cerebrovascular accidents (CVA). In this multicenter study, we explored the associations between clinical and imaging characteristics and outcomes of patients with LVNC, and externally validated the predictive value of myocardial thinning identified on cardiac magnetic resonance imaging (CMR) as previously described.

**Methods:** 214 adult patients (54% male, mean age 41±16 years) meeting the imaging criteria for LVNC were identified. Baseline clinical data, echocardiographic findings, and CMR characteristics were analyzed. Long-axis balanced steady-state free precession CMR sequences were used to assess myocardial thickness. Myocardial thinning was defined as a 50% or greater diameter reduction of the compacted myocardium compared to a contiguous segment. The primary endpoint was a composite of all-cause mortality, HF hospitalization, need of left ventricular assist device (LVAD) or heart transplant, cardiac resynchronization therapy (CRT), CVA/transient ischemic attacks (TIA), and VT and appropriate implantable cardioverter defibrillator (ICD) therapy.

**Results:** Focal myocardial thinning was observed in 42 patients (20%). Over a median follow-up period of 7 years (IQR, 4–10 years), 54 patients (24%) experienced a primary outcome (including all-cause mortality n=15 (7%), HF hospitalization/CRT n=33 (15%), CVA/TIA n=10 (5%), VT and appropriate ICD therapy n=14 (7%), LVAD/heart transplant n=3 (1.4%). Patients with myocardial thinning had more cumulative adverse events compared to those without myocardial thinning (chi-square = 29.516, log-rank < 0.001), even after matching for medical risk score. In a multivariate Cox regression model, myocardial thinning remained associated with outcomes: HR 3.052(95% CI: 1.569-5.937, p=0.001). When evaluating the incremental prognostic value of myocardial thinning over clinical and imaging characteristics, we observed that adding myocardial thinning added value to prediction models that included clinical and imaging features.

**Conclusions:** The presence of myocardial thinning is associated with adverse cardiovascular events in adult LVNC patients. Along with the medical risk score and other imaging variables, this feature enhances the risk stratification of individuals with LVNC. Incorporating myocardial thinning into medical risk assessments can improve the prediction and management of adverse outcomes in these patients.

## Introduction

Left ventricular non-compaction (LVNC), also known as left ventricular hypertrabeculation or noncompaction cardiomyopathy is a morphological abnormality characterized by prominent trabeculation of the left ventricle (LV), deep intertrabecular recesses that communicate with the ventricular cavity, and a thin compacted myocardial layer. Hypertrabeculation may occur in response to increased preload or afterload in patients with LV dysfunction and can coexist with various heart muscle disorders.^1^ Patients with LVNC may be asymptomatic or may present with a range of clinical manifestations, including life-threatening arrhythmias, heart failure (HF), systemic thromboembolic events, and sudden cardiac death. ^2^ Differentiating LVNC from increased trabeculation seen conditions, such as negative remodeling in different cardiomyopathies, athlete’s hearts, chronic volume, or pressure overload situations, poses a diagnostic challenge. Although many patients are first identified using echocardiography cardiac magnetic resonance imaging (CMR) has emerged as an important clinical tool to characterize patients with LVNC, because of its higher spatial resolution, better contrast between trabeculation and the blood pool, and no limitations in the acoustic window. CMR offers a more accurate and reliable evaluation of the extent of non-compacted myocardium than echocardiography and provides supplementary morphological information. Currently, the CMR diagnosis of LVNC using Petersen criterion is defined as a non-compacted to compacted myocardial ratio (NC/C ratio) greater than 2.3 in diastole, demonstrating a sensitivity of 86% and a specificity of 99%. ^3^ However, this criterion was based on a limited number of patients and was found to lack sensitivity in detecting prognostic indicators of adverse outcomes in patients with LVNC. In fact, the correlation between the extent and degree of hypertrabeculation and prognosis has to be established.(3, 4) More recently, a study by Ramchand et al. examined the role of myocardial thinning as measured by CMR and found that the risk of adverse clinical events increases in the presence of significant thinning of compacted myocardium, particularly in combination with elevated plasma natriuretic peptide levels. ^4^ The prognostic significance of myocardial thinning has been observed in studies across various conditions and imaging techniques. ^5^ However, data on the prognostic implications of CMR-based wall thickness measurements in cardiomyopathy remain limited.

The aim of our study was to investigate associations between clinical and imaging features and outcomes in patients with LVNC and specifically validate the predictive value of myocardial thinning identified on CMR in an external multi-center cohort. ^4^

## Methods

### Study population

For this observational, multicenter study patients with a diagnosis of LVNC were identified from research repositories at Stanford University and the Erasmus University Medical Center, A total of 214 adult patients ≥16 years of age who met Peterson criteria for LVNC on CMR performed between 2003 and 2023 at Stanford University (n=113 patients) or the Erasmus Medical Center (n=101 patients), were included in the study. Exclusion criteria included inadequate image quality, incomplete follow-up, and established diagnoses of another cardiomyopathy, such as ischemic or hypertrophic cardiomyopathy. Patients with a history of complex congenital heart disease (e.g. tetralogy of Fallot or transposition of the great arteries) were excluded. The study was approved by the institutional review boards at both centers (Stanford: Pro00042745; Erasmus MC Ethics Committee: MEC-2024-0155).

### CMR Assessment

CMR images were acquired using cine-balanced steady-state free precession sequences on 1.5-T or 3-T scanners (n=211 and n=3, respectively;). The slice thickness varied from 6 mm to 8 mm, the slice gap from 0 mm to 4 mm, and the median in-plane pixel spacing was 1.25 mm (interquartile range 0.70–1.44 mm). Biventricular volumes (end-diastolic and end-systolic), ejection fraction, and LV mass were calculated by manually tracing the endocardial and epicardial borders on steady-state free precession images. Late gadolinium enhancement (LGE) images were obtained in long-and short-axis orientations 15 to 20 minutes after injection of 0.2 mmol/kg of gadolinium chelate and qualitatively assessed.

LVNC was assessed using the Peterson criteria ^3^ as it has shown high inter- and intraobserver variability. ^6^ Following prior published CMR protocols, LVNC was qualitatively assessed on long-axis steady-state free precession cine images. The noncompacted and compacted layer was measured at the point of maximal trabecular thickness perpendicular to the border between the two layers. Papillary muscles and true apex were excluded from the measurements. A noncompacted/compacted >2.3 ratio in any segment during end-diastole established the presence of LVNC. ^3^

### Assessment of Myocardial Thinning

Myocardial thinning was assessed on long-axis CMR cine images following the methodology described by Ramchand et al. ^4^, and defined as a ≥50%-reduction of the compact myocardial wall thickness between the area of thinning and the adjacent myocardium within the same image (Figure 1).

**Figure 1.**
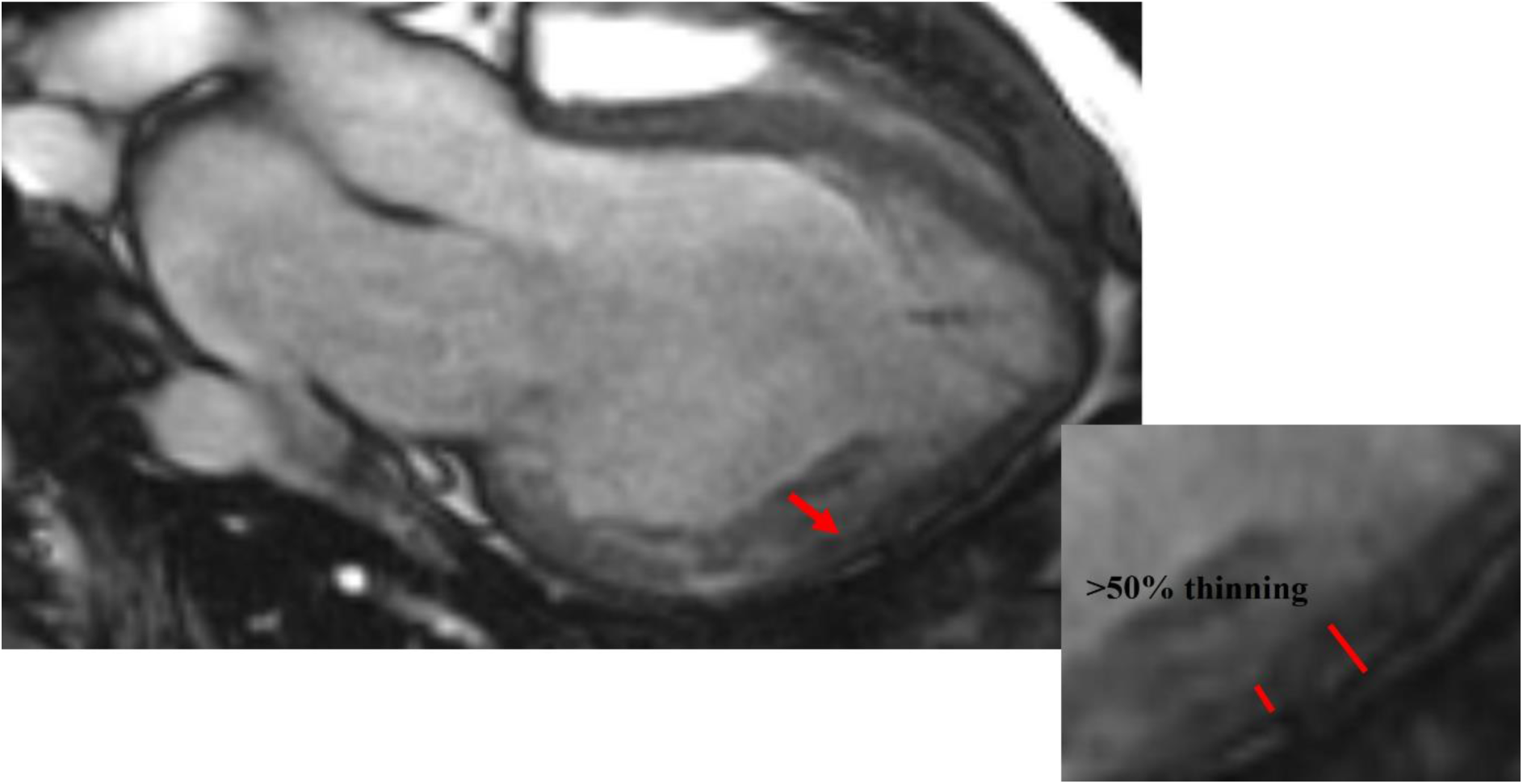
Abrupt myocardial thinning. Basal to mid-inferolateral segments thinning. Red arrows and lines highlight abrupt myocardial thinning, defined by the compacted myocardium’s thinning by ≥50% compared with a contiguous myocardial segment.

### Follow-up and endpoint

The composite endpoint of this study was defined as all-cause mortality, HF hospitalization, cardiac resynchronization therapy (CRT), CVA/transient ischemic attacks (TIA), and sustained ventricular tachyarrhythmia (VT), and appropriate implantable cardioverter defibrillator (ICD) therapy. The duration of follow-up ranged from the CMR exam to the first event/last office follow-up. Endpoint data were obtained by reviewing the electronic information system and retrieval of survival status through the medical records.

### Statistical Analysis

Continuous variables are presented as mean ± SD if normally distributed or as median and interquartile range otherwise. Categorical variables are presented as frequencies and percentages. One-way analysis of variance and the Mann-Whitney test were used for normally distributed and skewed variables, respectively, whereas the χ^2^ test was used to compare categorical variables. Propensity matching score was used to match groups with and without myocardial thinning based on medical risk score (combining variables such as: sex, age, diabetes mellitus, hypertension, atrial fibrillation (AF), VT, hyperlipidemia, HF, stroke, CVA/TIA, left and right bundle branch block (LBBB/RBBB), and family history of LVNC or related cardiomyopathies). The matching tolerance was set at 0.01. To assess the hazard ratio (HR) change for adverse outcomes across a range of compacted myocardial thinning values, a spline curve analysis was performed.

Cumulative event rates were analyzed using the Kaplan-Meier survival method, stratifying patients into two groups based on LV wall thinning of ≥50% compared to <50%, and comparisons were made using the log-rank test. The association between myocardial thinning and cardiovascular events was evaluated using uni- and multivariate Cox regression analyses.

The level of significance for variables to be included in the multivariable analysis was set at *P* < .05 and *P<0.001.* Hazard ratios (HRs) and 95% confidence intervals (Cis) are presented. To assess the incremental value of myocardial thinning, we compared the χ^2^ values of different multivariate Cox regression models that included clinical and imaging variables previously shown to have prognostic value.

The medical risk score indicating relative risk of major adverse cardiovascular events (MACE) was previously established by Ramchand et al. ^4^ The univariate relationship between each medical risk factor - sex, age, diabetes mellitus, hypertension, atrial arrhythmias, VT, hyperlipidemia, HF, CVA/TIA, systemic embolization, LBBB, RBBB, and family history of LVNC – and the risk of MACE was assessed using Cox proportional hazards regression models. Consequently, significant variables were selected to build a risk score. The risk was calculated by multiplying the regression coefficient of each significant risk factor by the value of that variable.

Statistical analysis was performed using SPSS for Windows version 29.0 (IBM, Armonk, NY) and in R environment 3.6.4 (R Foundation for Statistical Computing). A two-tailed *P* value < .05 was considered to indicate statistical significance.

## Results

### Baseline clinical characteristics

A total of 214 patients with LVNC were included (46% women, mean age 41±16 years). Baseline clinical characteristics, medications, imaging, and biomarkers are listed in Tables 1, and 2. The cohort exhibited a high prevalence of HF, followed by VT, AF, and CVA/TIA Cardiovascular risk factors and medications are listed in Table 1. A family history of LVNC or other cardiomyopathy phenotypes was documented in one-third of the study population (Table 2). Patients with compact myocardial thinning more frequently had a history of CVA/TIA and VT (Table 1).

**Table 1:** Baseline clinical characteristics of the total study population divided by the presence of compact myocardial thinning.

| Baseline characteristics | Overall<br>n=214 | Myocardial<br>thinning present<br>n=42 | Myocardial<br>thinning absent<br>n=172 | p-value |
| --- | --- | --- | --- | --- |
| **Clinical Characteristics** |  |  |  |  |
| Age, y | 41±16 | 42±16 | 41±16 | 0.691 |
| Women, n (%) | 98 (46) | 23 (55) | 75 (44) | 0.130 |
| Diabetes mellitus, n (%) | 14 (7) | 2 (5) | 12 (7) | 0.457 |
| Hypertension, n (%) | 41 (19) | 7 (17) | 34 (20) | 0.416 |
| Hyperlipidemia, n (%) | 36 (17) | 6 (14) | 30 (17) | 0.409 |
| Heart failure, n (%) | 83 (39) | 20 (47) | 63 (30) | 0.129 |
| Atrial fibrillation, n (%) | 35 (16) | 12 (29) | 23 (13) | **0.019** |
| CVA/TIA, n (%) | 14 (7) | 7 (17) | 7 (4) | **0.008** |
| Ventricular arrhythmias, n (%) | 46 (21) | 18 (43) | 28 (16) | **<0.001** |
| FH of LVNC or another phenotype, n (%) | 59(29) | 10 (26) | 53 (31) | 0.318 |
| **Medical Therapy** |  |  |  |  |
| ACEi, n (%) | 69 (32) | 16 (38) | 53 (31) | 0.234 |
| ARB/ARNi, n (%) | 46 (21) | 16 (39) | 30 (17) | **0.005** |
| B-blocker, n (%) | 128 (60) | 31 (76) | 96 (56) | **0.011** |
| Aldosterone antagonist, n (%) | 36 (17) | 11 (26) | 25 (15) | 0.063 |
| Aspirin, n (%) | 46 (22) | 7 (17) | 39 (23) | 0.266 |
| NOAC, n (%) | 43 (20) | 11 (26) | 32 (19) | 0.191 |

ACEi, angiotensin converting enzyme inhibitor; ARB/ARNi, angiotensin receptor blocker/angiotensin receptor neprilizin inhibitor; CVA, cerebrovascular accident; FH, family history; LVNC, left ventricular non-compaction; NOAC, novel oral anticoagulant; TIA, transient ischemic attack

**Table 2:** Baseline CMR imaging, ECG characteristics, and blood biomarkers of the total study population divided by the presence of compact myocardial thinning.

| **Baseline characteristics**<br>n=214 | Overall<br>n=214 | Myocardial<br>thinning<br>present<br>n=42 | Myocardial<br>thinning<br>absent<br>n=172 | p value |
| --- | --- | --- | --- | --- |
| **Imaging (CMR)/ characteristics** |  |  |  |  |
| LVEDVi, mL/m<sup>2</sup> | 116±43 | 134±38 | 112±43 | **0.003** |
| LVESVi, mL/m<sup>2</sup> | 70±36 | 90±41 | 65±33 | **<0.001** |
| LVSVi, mL/m<sup>2</sup> | 49±15 | 44±12 | 50±15 | **0.016** |
| LVEF, % | 44±13 | 36±13 | 46±12 | **<0.001** |
| LVMi, g/m<sup>2</sup> | 64 (54-74) | 69 (64-74) | 60 (52-73) | 0.429 |
| NC/C ratio | 2.7 (2.6-2.8) | 3.0 (2.6-3.5) | 2.7 (2.5-3.2) | 0.084 |
| Mean number of trabeculated segments | 3.12±0.94 | 3.11±0.93 | 3.14±1.00 | 0.331 |
| Late gadolinium enhancement, n (%) * | 46 (25) | 18 (49) | 28 (19) | **<0.001** |
| RV hyper trabeculation, n (%) | 55 (29) | 18 (47) | 37 (24) | 0.164 |
| Abnormal RV size and function, n (%) | 54 (29) | 17 (47) | 37 (25) | **0.010** |
| Elevated BNP/NTproBNP, n (%) * | 27 (23) | 14 (50) | 13 (14) | **<0.001** |
| **ECG characteristics** |  |  |  |  |
| Left Bundle branch block | 22 (10) | 3 (7) | 19 (11) | 0.336 |
| Right bundle branch lock | 8 (4) | 2 (5) | 6 (4) | 0.488 |
| Any bundle branch block | 29 (14) | 5 (12) | 24 (14) | 0.477 |

BNP/ NTproBNP, brain natriuretic peptide/ N-terminal prohormone of brain natriuretic peptide; CMR, cardiac magnetic resonance imaging; ECG, electrocardiography; LVEDVi, left ventricular end-diastolic volume indexed; LVEF, left ventricular ejection fraction; LVESVi, left ventricular end-systolic volume indexed; LVMi, left ventricular mass index; LVSVi, left ventricular stroke volume indexed; NC/C, noncompacted/compacted ratio; RV, Right ventricle.
\*Late gadolinium enhancement is evaluated in n=187 patients
\*BNP/NTproBNP is evaluated in n=119 patients

The study populations from Stanford University and Erasmus Medical Center Rotterdam exhibited similar clinical characteristics, except for HF and VT prevalence, which was higher in the Stanford University cohort (Supplemental Table 1).

### Cardiac Magnetic Resonance

On average, LVNC patients in this cohort had large LV end-systolic (LVESVi) and end-diastolic indexed volumes (LVEDVi), a reduced LV ejection fraction (LVEF), and a high left ventricular mass index (LVMi). Additionally, 29% were found to have abnormal right ventricular (RV) size or function (Table 2). LGE imaging was performed in 187 patients and showed myocardial enhancement in 46 (25%). The mean number of myocardial segments per patient meeting the Petersen criteria was 3.1, most often involving the apical segments and basal to mid-ventricular lateral segments, followed by inferior and anterior segments. The least frequently affected were the basal to mid-ventricular septal segments. Compact myocardial thinning was observed in 42 patients (20%). Patients with myocardial thinning had larger LV volumes, lower LVEF, more frequent RV abnormalities, and a higher prevalence of LGE compared to patients without myocardial thinning (Table 2).

### Outcomes in patients with and without myocardial thinning

After a median follow-up of 7 years (IQR, 4-10), adverse events occurred in 54 (25%) patients, including all-cause mortality in 15 (7%), HF hospitalization/CRT in 33 (15%), CVA/TIA in 10 (5%), and VT in 14 (7%) (Figure 2). Patients with myocardial thinning exhibited a higher prevalence of cumulative events compared to those without myocardial thinning (59% vs.17%; chi-square = 29.516, log-rank < 0.001; Figures 2, 3 A). When using propensity score matching to match patients with and without myocardial thinning according to medical risk score (n=68 patients selected), patients with myocardial thinning still experienced higher cumulative rates of cardiovascular events/death as compared to patients without myocardial thinning (χ2 =6.396; log-rank= 0.011, Figure 3B).

**Figure 2.**
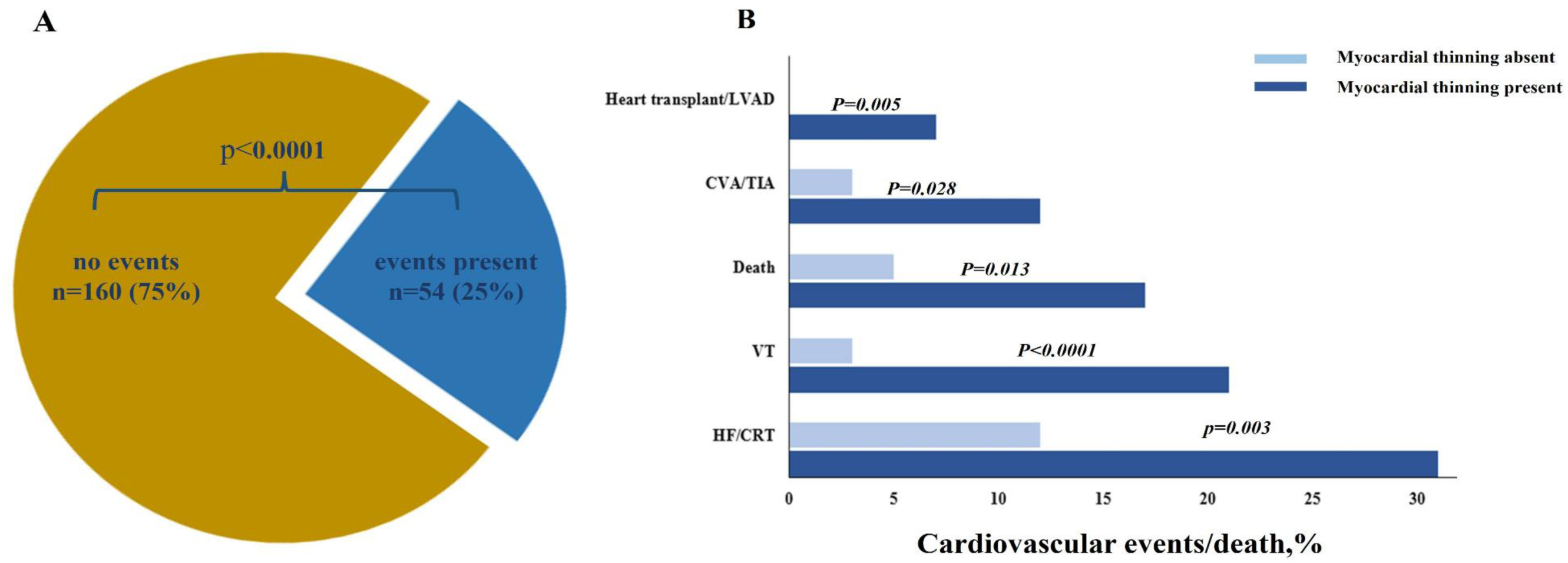
Prevalence of adverse cardiovascular events/death in the overall population (A) and divided by the presence of compact myocardial thinning (B) CRT, cardiac resynchronization therapy; CVA, cerebrovascular accident; HF, heart failure; LVAD, left ventricular assist device; TIA, transient ischemic attack; VT, ventricular arrhythmia.

**Figure 3.**
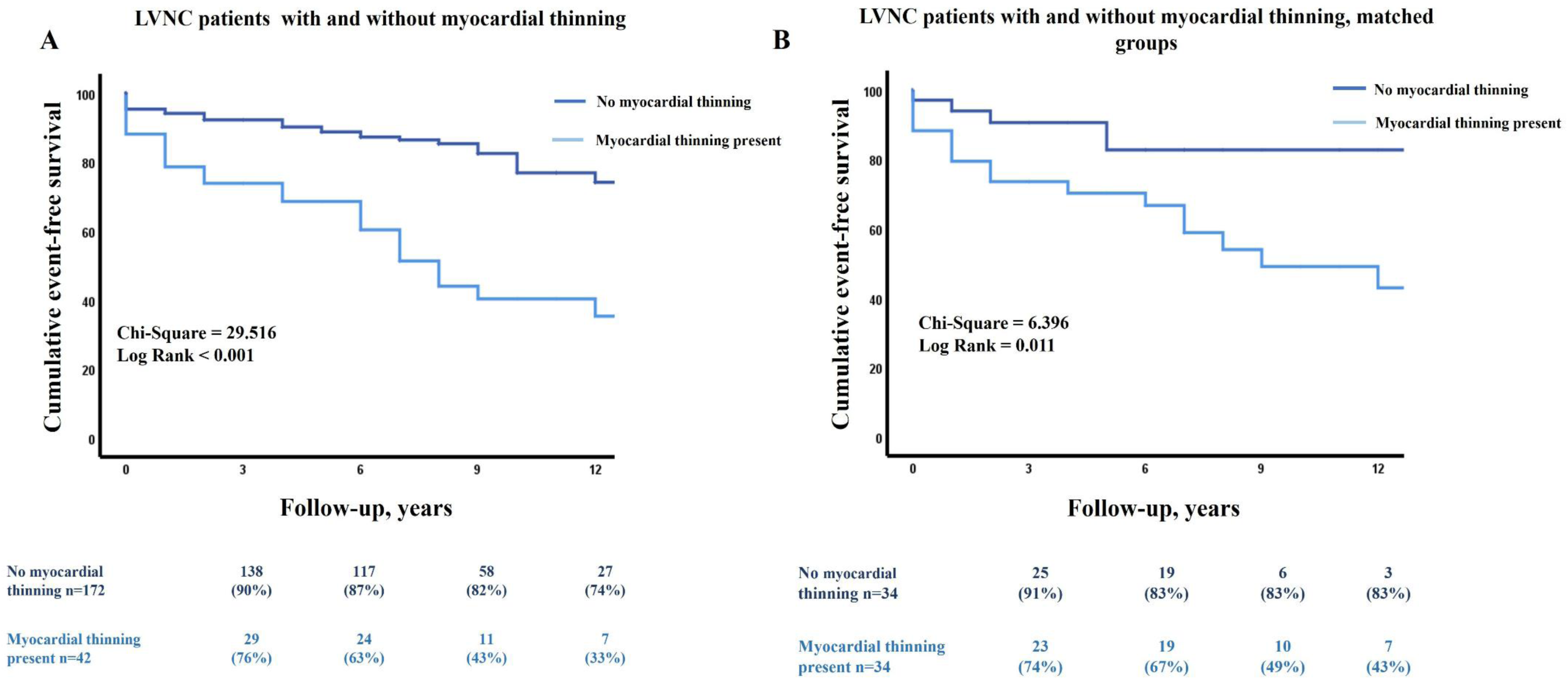
Survival analysis in patients with LVNC with and without compact myocardial thinning (A) and after adjusting for medical risk score (B). propensity score matching was applied to match the groups for sex, age, diabetes mellitus, hypertension, AF, VT, hyperlipidemia, HF, CVA/TIA, LBBB, RBBB, family history of LVNC, n=82 patients selected. LVNC, left ventricular non-compaction.

Multivariable Cox proportional hazards regression models were constructed to evaluate the independent associations with outcomes incorporating various imaging and clinical characteristics (Table 3). In the univariate analysis, several parameters were significantly associated with adverse events: age, AF, VT, HF, LBBB, family history of LVNC or related phenotypes, BNP/NTproBNP, myocardial thinning, LGE, and LVESVi.

**Table 3:** Univariate and Multivariate cox regression analysis.

| Clinical and imaging characteristics | Univariate analysis |  | Multivariate analysis (Model I) |  | Multivariate analysis (Model II) |  |
| --- | --- | --- | --- | --- | --- | --- |
|  | HR (95%CI) | *P* | HR (95%CI) | *P* | HR (95%CI) | *P* |
| Women | 0.550 (0.311-0.971) | **0.039** | 1.182 (0.403-3.464) | 0.761 |  |  |
| Age | 1.035 (1.018-1.053) | **<0.001** | 1.008 (0.979-1.039) | 0.582 | 1.01(0.996-1.042) | 0.105 |
| Diabetes mellitus | 2.081 (0.938-4.614) | 0.073 |  |  |  |  |
| Hypertension | 1.697 (0.946-3.046) | 0.077 |  |  |  |  |
| Atrial Fibrillation | 3.240 (1.820-5.767) | **<0.001** | 3.036 (1.045-8.814) | **0.041** | 1.926 (0.913-4.059) | 0.085 |
| Ventricular arrhythmia | 2.529 (1.448-4.417) | **<0.001** | 1.162 (0.440-3.066) | 0.762 | 2.016 (1.027-3.958) | **0.042** |
| Hyperlipidemia | 1.653 (0.906-3.017) | 0.101 |  |  |  |  |
| Heart failure | 2.750 (1.588-4.760) | **<0.001** | 0.649 (0.227-1.854) | 0.419 | 1.461 (0.745-2.867) | 0.270 |
| CVA/TIA | 1.726 (0.685-4.347) | 0.247 |  |  |  |  |
| Left bundle branch block | 3.154 (1.710-5.816) | **<0.001** | 5.902 (1.262-27.593) | **0.024** | 2.794 (1.311-5.954) | **0.008** |
| Right bundle branch block | 0.744 (0.181-3.064) | 0.682 |  |  |  |  |
| FH of LVNC or related phenotypes | 0.299 (0.133-0.673) | **0.004** | 0.453 (0.105-1.959) | 0.289 |  |  |
| Myocardial thinning | 3.889 (2.274-6.651) | **<0.001** | 4.164 (1.192-14.543) | **0.025** | 3.052 (1.569-5.937) | **0.001** |
| Late gadolinium enhancement | 4.042 (2.232-7.322) | **<0.001** | 2.260 (0.689-7.408) | 0.178 | 1.636 (0.771-3.468) | 0.200 |
| LVESVi mL/m<sup>2</sup> | 1.019 (1.013-1.025) | **<0.001** | 1.008 (0.995-1.022) | 0.240 | 1.015 (1.005-1.022) | **0.003** |
| NTproBNP/BNP | 3.075 (1.534-6.163) | **0.002** | 0.999 (0.302-3.303) | 0.999 |  |  |

CVA, cerebrovascular accident; LVESVi, left ventricular end-systolic volume indexed; BNP/ NTproBNP, brain natriuretic peptide/ N-terminal prohormone of brain natriuretic peptide; TIA, transient ischemic attack

Myocardial thinning was associated with outcomes in multivariate models after adjusting for significant clinical and imaging characteristics (**model I:** HR 4.164; 95% CI 1.192 to 14.543; p =0.025, **model II**: HR 3.052; 95% CI 1.569 to 5.937; p =0.001; Table 3) alongside with VT, LBBB, and LVESVi. After adjusted for the medical risk score alone, myocardial thinning remained significantly associated with outcomes (HR 3.517; 95% CI 2.051 to 6.032; p<0.001; Table 4).

**Table 4:**
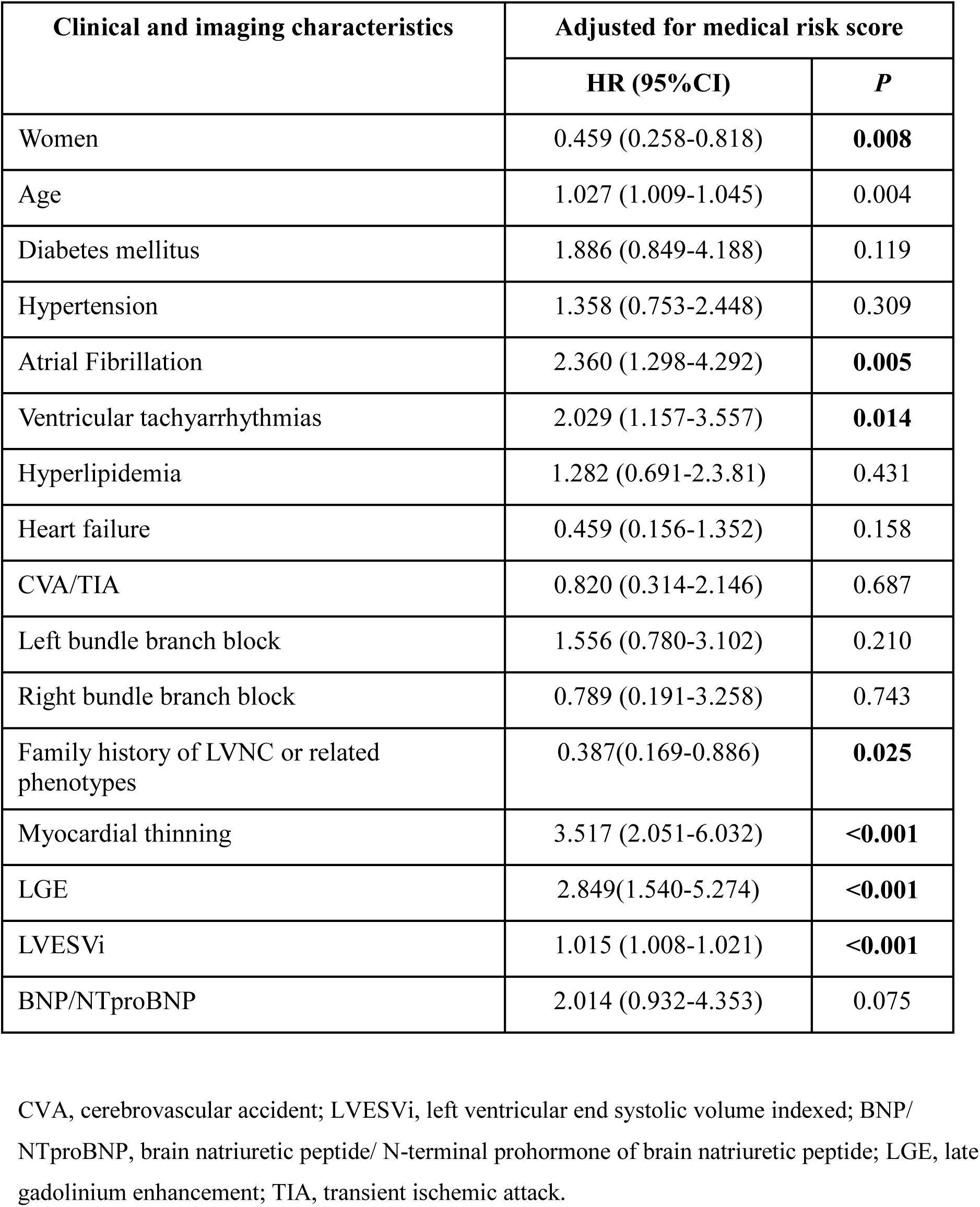
Clinical and imaging characteristics adjusted for medical risk score.

| Clinical and imaging characteristics | Adjusted for medical risk score |  |
| --- | --- | --- |
|  | HR (95%CI) | *P* |
| Women | 0.459 (0.258-0.818) | **0.008** |
| Age | 1.027 (1.009-1.045) | 0.004 |
| Diabetes mellitus | 1.886 (0.849-4.188) | 0.119 |
| Hypertension | 1.358 (0.753-2.448) | 0.309 |
| Atrial Fibrillation | 2.360 (1.298-4.292) | **0.005** |
| Ventricular tachyarrhythmias | 2.029 (1.157-3.557) | **0.014** |
| Hyperlipidemia | 1.282 (0.691-2.3.81) | 0.431 |
| Heart failure | 0.459 (0.156-1.352) | 0.158 |
| CVA/TIA | 0.820 (0.314-2.146) | 0.687 |
| Left bundle branch block | 1.556 (0.780-3.102) | 0.210 |
| Right bundle branch block | 0.789 (0.191-3.258) | 0.743 |
| Family history of LVNC or related phenotypes | 0.387(0.169-0.886) | **0.025** |
| Myocardial thinning | 3.517 (2.051-6.032) | **<0.001** |
| LGE | 2.849(1.540-5.274) | **<0.001** |
| LVESVi | 1.015 (1.008-1.021) | **<0.001** |
| BNP/NTproBNP | 2.014 (0.932-4.353) | 0.075 |

CVA, cerebrovascular accident; LVESVi, left ventricular end systolic volume indexed; BNP/NTproBNP, brain natriuretic peptide/ N-terminal prohormone of brain natriuretic peptide; LGE, late gadolinium enhancement; TIA, transient ischemic attack.

When evaluating the incremental prognostic value of myocardial thinning over clinical and imaging characteristics, we observed that incorporating myocardial thinning improved the predictive performance of models that included clinical features (age, HF, AF, LBBB or RBBB, BNP/NTproBNP) and traditional imaging characteristics (LVESVi, LGE, basal to mid inferior segment hypertrabeculation) (Figure 4).

**Figure 4.**
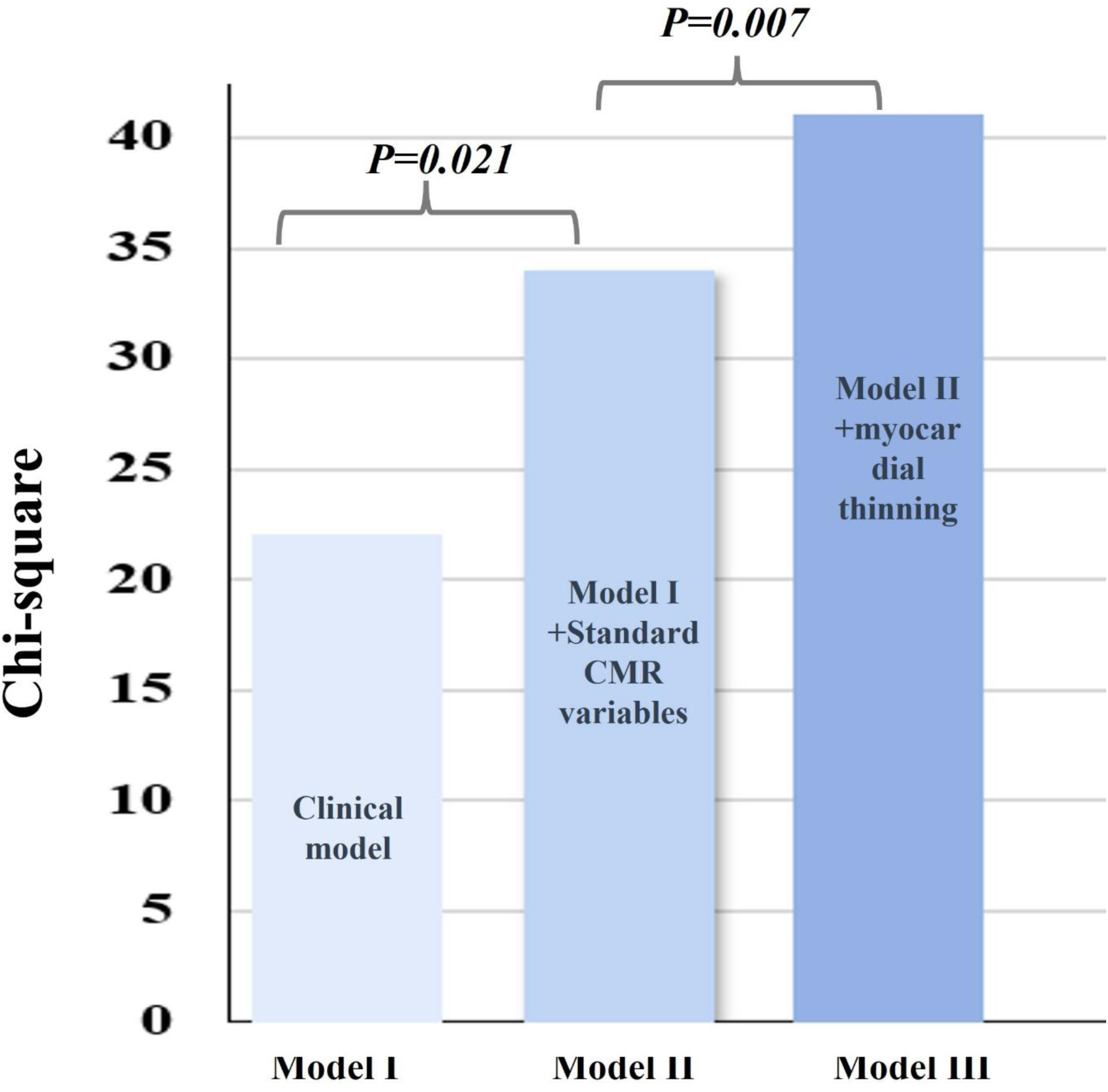
Prognostic value of compacted myocardial thinning calculated with chi-square over clinical variables and imaging characteristics. **Model I** includes age, heart failure, atrial fibrillation, any bundle branch block, and BNP/NTproBNP **Model II** added LGE, LVESVi, basal to mid inferior segment hypertrabeculation, and **Model III** added focal myocardial thinning. BNP/NTproBNP, brain natriuretic peptide/ N-terminal prohormone of brain natriuretic peptide; CMR, cardiac magnetic resonance imaging; LGE, late gadolinium enhancement; LVESVi, left ventricular end-systolic volume index.

Additionally, to investigate the relationship between myocardial thinning and adverse outcomes, a spline curve analysis was performed (Figure 5). After an initial slow rise in HR, there was an increase in the HR of adverse outcomes for myocardial thinning starting at 40% and increasing gradually, indicating that when myocardial thinning exceeds 40%, the risk of adverse outcomes escalates significantly.

**Figure 5.**
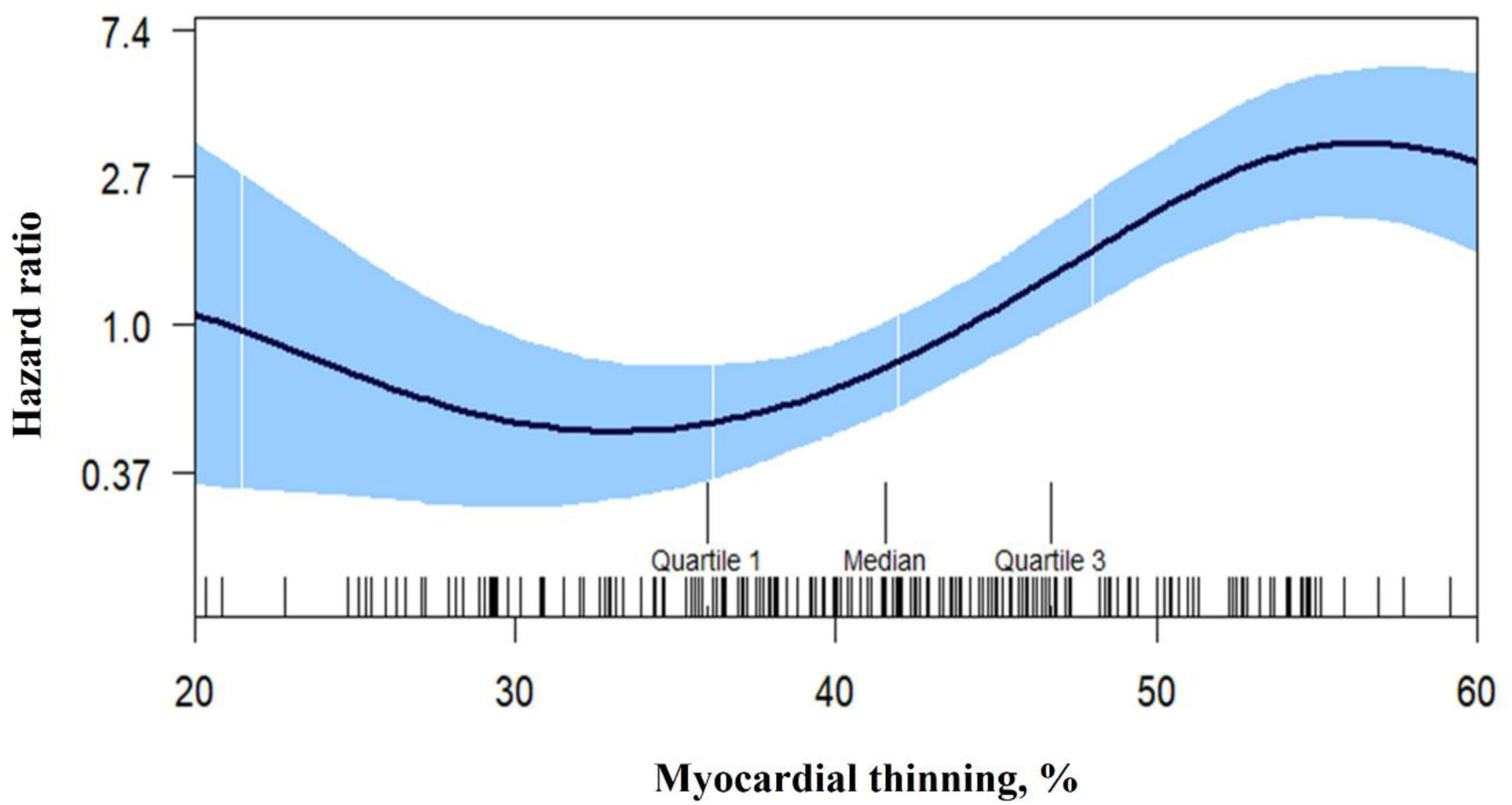
Spline curve for adverse outcomes according to compacted myocardium thinning. The curve represents the hazard ratio change for adverse events with overlaid 95% confidence intervals (light blue) across a range of myocardial thinning.

## Discussion

Our multicenter study investigating clinical and imaging predictors of outcomes in patients with LVNC confirms prior observations that thinning of the compact myocardium on CMR is associated with adverse outcomes, alongside factors such as VT, LVESVi, and LBBB. In our cohort, patients exhibiting focal myocardial thinning ≥50% compared to adjacent myocardial segments experienced 3 times worse outcomes than those without thinning (HR 3.052; 95% CI 1.569 to 5.937; p =0.001).

The clinical presentation of LVNC is highly variable, it can manifest at any age and range from asymptomatic to end-stage heart failure, and the condition has been associated with life-threatening arrhythmias, sudden cardiac death, or thromboembolic events. ^7–9^ While the diagnosis of LVNC has been primarily based on identifying and quantifying abnormal trabeculations, in this study, we aimed to determine the prognostic significance of the compacted wall thickness compared to established clinical and imaging predictors of outcome.

### Value of compact myocardial thinning in the prediction of outcomes

The diagnosis of LVNC is mostly based on non-invasive imaging studies, with transthoracic echocardiography (TTE) and CMR being the most widely utilized methods. TTE is often the initial diagnostic approach due to its availability and lower cost. The major TTE criteria for diagnosing LVNC rely on the ratio of the thickness of the non-compacted layer to that of the compacted layer. TTE also provides valuable insights into the function and structure of the left ventricle. ^10^

However, echocardiographic parameters often lack the sensitivity needed to differentiate normal from potentially pathological hypertrabeculation. To enhance the diagnostic accuracy of LVNC, advanced echocardiographic techniques such as strain and strain rate imaging are increasingly utilized. ^11^ Conversely, CMR is becoming increasingly valuable for diagnosing and monitoring LVNC, as it provides crucial insights into both the structure and function of the left ventricle, along with important prognostic information. ^12^

The most common diagnostic criteria for LVNC using CMR are also based on the ratio of the thickness of the non-compacted layer to that of the compacted layer, with a threshold of greater than 2.3 at the end of diastole, as suggested by Petersen et al^3^ CMR has demonstrated that in LVNC the compact layer is often abnormally thin, particularly at the apex, which can be mistaken for apical aneurysms. The prognostic implications of myocardial thinning on CMR have yet to be thoroughly studied.

Lazzari et al in 33 patients with isolated LVNC observed that a thinned compact layer of mid-ventricular segments of the LV free wall was associated with reduced systolic function.^13^ A study by Jang et al., demonstrated that slower conduction velocity was observed in the presence of myocardial wall thinning in a swine model of healed left ventricular infarction during CMR evaluation. ^14^ Emerging data from cardiac CT studies also suggest that severe wall thinning found in ischemic cardiomyopathy and post-myocarditis is a useful tool to identify VT substrate and helpful for understanding the mechanisms of the location of the VT substrate domain.^15^ study by Galand et al demonstrated that left ventricular wall thickness measured on cardiac CT could be associated with the response to CRT therapy. ^5^ Additionally, the study by Kaminaga et al. identified LGE, focal wall thinning, and fatty components as abnormal findings in patients with dilated cardiomyopathy using ultra-fast CT imaging.^16^

In this context, the study by Ramchand et al. explored the prognostic significance of abrupt myocardial thinning in patients with LVNC and found a notable association with outcomes across various clinical models.^4^ Our study confirmed these findings by employing similar models, proposed in their research, and additionally demonstrated the association of myocardial thinning with the outcomes as well as with medical risk score, LVESVi, and LGE (Supplemental Table 2). These associations were observed across various clinical models that included key imaging and clinical variables based on our results.

### Clinical predictors of cardiovascular outcomes in LVNC patients

When evaluating the predictive value of various clinical characteristics, we observed that in multivariate analysis, VT, LBBB, and LVESVi were clinical variables significantly associated with outcomes, together with myocardial thinning. Our findings align with prior studies that have demonstrated a significant association between age and AF with CVA/TIA in LVNC patients. ^4,17^ The prevalence of AF in patients with LVNC varies from 1 to 29% ^18,19^. The pathophysiology of AF in LVNC is largely unknown. However, underlying myopathy, LA dilation, and/or ion channel changes are the leading suspected causes of AF in adult LVNC.^20,21^ Studies by Stollberger et al. found that older age, exertional dyspnea, diabetes mellitus, and heart failure were more common in LVNC patients with AF, therefore, our observations may reflect more prevalent cardiac and extracardiac co-morbidity in patients with AF. ^22^

Our study confirms prior observations that LBBB is associated with adverse outcome^23^ Abnormal trabecular meshwork (associated with affected embryogenesis and unmatured conduction system) in LVNC may lead to conduction abnormalities and impede electrical conduction pathways. ^24^ The trabeculated, non-compacted myocardium can already create areas of electrical heterogeneity, leading to abnormal conduction patterns and arrhythmias including ventricular tachycardia and atrial fibrillation.^25,26^ Ischemic events, the presence of fibrosis, and genetic predispositions are other contributing factors for LBBB. ^23,26–33^ Our study confirms the findings of prior studies that LGE on CMR is associated with adverse outcomes. ^1,34,35^

In our study, myocardial thinning was associated with outcomes in each of the multivariate models, even after adjusting for above mentioned significant clinical and imaging characteristics. These results are in line with the findings of Ramchand et al, ^4^ which also demonstrated the association of myocardial thinning with the outcomes in LVNC patients across different regression models. Notably, the baseline clinical and imaging characteristics of our population closely resembled those reported by Ramchand et al., with the exception of a lower prevalence of hypertension and hyperlipidemia.

LVESVi was another imaging variable associated with the outcomes in our multivariate models. Studies have shown, that in patients with LVNC, alongside adverse remodeling, LVESVi can reflect changes in chamber geometry and function over time. Factors like age at initial presentation, and the presence of cardiovascular conditions such as AF, HF, CVA/TIA, and VT, as well as increased LV volumes and dimensions, can influence prognosis. We adopted the medical risk score developed by Ramchand et al which included important clinical variables such as sex, age, diabetes mellitus, hypertension, AF, VT, hyperlipidemia, HF, stroke, systemic embolization, LBBB, RBBB, family history of LVNC and validated these findings in our multivariate models. ^4^ (Supplemental table 2).

## Limitations of the study

The strength of our study lies in its retrospective, observational design across two centers, which helps validate the findings from the Cleveland Clinic. However, there are several limitations to consider. First, the total area of myocardial thinning was not measured in this study. It is expected that focal myocardial thinning reflects the overall pattern of myocardial involvement, with more extensive thinning likely associated with worsening left ventricular function and/or poorer clinical outcomes.

Additionally, late gadolinium enhancement imaging was not performed in all patients; it was available in 87% of cases across the two centers. Furthermore, our study did not find evidence to support an association between BNP/NT-proBNP levels and outcomes in patients with LVNC. It’s important to note, that baseline biomarker measurements, which were taken near the time of CMR evaluation, were only available for 56% of the population. This limited data availability restricts our ability to draw definitive conclusions about the predictive value of these biomarkers. Clinical and imaging characteristics and adverse cardiovascular outcomes suggest possible associations; however, they do not establish a causal relationship.

## Conclusions

Our study demonstrates that while traditional diagnostic criteria for LVNC focus on trabeculations, the analysis of myocardial thinning through advanced imaging techniques like CMR provides valuable insights into patient outcomes. Furthermore, our study performed in the multi-center setting underscores the importance of clinical factors such as VT, LVESVi and LBBB as significant predictors of outcomes alongside myocardial thinning. This underlines the necessity for a comprehensive approach to the diagnosis and monitoring of LVNC patients.

## Data Availability

Data will be available upon request

## Disclosures

**Tea Gegenava**-None; **Koen Nieman**-Dr. Nieman acknowledges support from the NIH (NIH R01- HL141712; NIH R01 - HL146754); unrestricted institutional research support from Siemens Healthineers, Novartis; consulting for Cleerly, Artrya, RapidAI and Novartis; equity in Lumen Therapeutics, all unrelated to this work**. A. Hirsch** received a research grant and consultancy fees from GE Healthcare and speaker fees from GE Healthcare, Bayer, and Bristol Myers Squibb. He is also a member of the medical advisory board of Medis Medical Imaging Systems and was MRI corelab supervisor of Cardialysis BV until 2022.

## Acknowledgments

**Ashish Manohar**-this work was done during the term of an Award from the American Heart Association (AHA 24POST1187968).

## Notes

### Competing Interest Statement

The authors have declared no competing interest.

### Author Declarations

The study was approved by the institutional review boards at both centers (Stanford: Pro00042745; Erasmus MC Ethics Committee: MEC-2024-0155)

